# Longitudinal phase 2 clinical trials of live, attenuated tularemia vaccine in otherwise healthy research laboratory workers operating in containment laboratories

**DOI:** 10.1101/2023.01.09.23284371

**Authors:** David L. Saunders, Benjamin C. Pierson, Jeannine Haller, Sarah Norris, Anthony P. Cardile, Ronald B. Reisler, Arthur C. Okwesili, Ellen Boudreau, Janice Rusnak, Denise K. Danner, Bret K. Purcell, James F. Barth, Erin L. Tompkins, Isaac L. Downs, Dani Liggett, Patricia Pettit, Tami Pratt, Mark Goldberg, Mark G. Kortepeter, Fernando B. Guerena, John W. Aldis, Maryam Keshtkar, Phillip R. Pittman

## Abstract

**Background:** Tularemia is a bacterial disease caused by the intracellular bacterium *Francisella tularensis* (*F. tularensis* or *Ft*). It has been weaponized historically by multiple state actors due to its low infectious aerosol dose, high morbidity and high mortality rate of the pneumonic form. The US Army developed the attenuated Live Vaccine Strain (LVS) from stocks provided by the former Soviet Union in the 1950s. The vaccine has proven to be safe and immunogenic over the ensuing decades in numerous clinical trials and animal as well as human challenge studies. Despite the threat, there are no FDA-approved vaccines nor clinical stage candidates against tularemia. LVS remains unlicensed due to instability in culture and the potential for reversion to the wild-type pathogen. We report here two sequential LVS trials in at-risk laboratory personnel working on tularemia in bio-containment.

**Methods:** Volunteers received a single dose of the Live Vaccine Strain (LVS) live, attenuated tularemia vaccine by scarification under 2 FDA-regulated non-randomized, single-arm protocols (IND 157). Positive immunization was based on local scarification site ‘take reaction’, and either a >1:20 tularemia antigen microagglutination (MA) titer (protocol FY03-24; 2004-8) or greater than 4-fold rise in MA titer (protocol FY07-15; 2009-2017). Those still negative by week 4 were offered a second dose.

**Results:** The LVS vaccine was safe, well tolerated and highly immunogenic. Between the two studies, all recipients (100%) had positive ‘take reactions’, with 95.5% of those in study FY03-24 having a positive response following initial vaccination. All but 3 subjects (98%) in protocol FY03-24 had positive MA titer results defined as >1:20, most within 28-35 days. In protocol FY07-15, 95% of subjects had a 4-fold or greater rise in MA titer, the primary immunogenicity endpoint for that study.

**Conclusions:** LVS vaccine administered to laboratory workers at risk for tularemia exposure over a 12 year period was safe and highly immunogenic. Findings were in line with more than 4 decades of prior similar results. Response rates remained robust despite the vaccine lots employed having been manufactured 2-3 decades prior to the present studies. In the absence of a commercial development effort, or another tularemia vaccine in clinical development, a vaccine protocol under investigational new drug (IND) application could be considered based on the large body of favorable data for this vaccine. The results as well as historical comparator data presented here should serve as a benchmark for future studies.

## Introduction

*Francisella tularensis* infects vertebrate and invertebrate animal hosts causing the disease tularemia (1). Transmission to mammals in nature occurs largely through multiple vectors including ticks, mosquitos and biting flies, and contact with remains of infected animals. Clinical presentations vary with skin, eye, lymphatic, gastrointestinal and pharyngeal involvement common following a 3-5 day incubation period (2, 3). While less commonly fatal, even naturally transmitted disease can last several weeks followed in some cases by months of chronic fatigue (4). An extremely low infectious dose (10-50 organisms via aerosol), ease of aerosolization, and greater virulence when inhaled make it an ideal bioweapon (5) capable of incapacitating and killing large enemy formations. Aerosolized tularemia may be fatal in 60% of untreated individuals (6). Tularemia has been classified on the U.S. Federal Select Agents and Toxins List as a Tier 1, CDC Category A biological agent, and Risk Group 3 pathogen capable of causing serious disease (7).

Tularemia can cause infection by multiple routes, causing various cutaneous, glandular and ocular syndromes. The more serious pneumonic form of the disease occurs commonly in laboratory workers exposed by the aerosol or oral routes. Laboratory-acquired infection with *F. tularensis* is a well-recognized hazard of *Ft* biomedical research, with >200 cases described in the American medical literature (8, 9). Pneumonic disease is the syndrome most likely to be encountered in a deliberate attack. It is characterized by fever, malaise, pneumonic infiltrates on x-ray, severe prostration, and diarrhea lasting 3-6 weeks. Survivors may suffer chronic fatigue for months. Individuals suffering pneumonia or pleuritic infection face a poor prognosis. Death is most common with the pneumonic form. Tularemia contracted from zoonotic sources can be treated with antibiotics including streptomycin, gentamicin, doxycycline, and ciprofloxacin. Recommended treatment duration is 10-21 days depending on the stage of illness and medication used, though it is unclear how well conventional antibiotics would perform in a deliberate aerosol attack. Substantial insights on its potential for weaponization were gained in part by 40 challenge experiments on healthy volunteers, participants in ‘Operation White Coat’, exposed to various doses over 10 years starting in 1958 (10). There is a clear infectious dose-response relationship with reduced time of onset and more severe disease occurring at higher exposures. Aerosolized tularemia may relapse if not treated with 10-14 days of sufficient antibiotic therapy equivalent to > 2 grams per day tetracycline (10). Though tetracycline itself is no longer recommended, doxycycline continues to be a treatment option as well as fluoroquinolones and aminoglycosides (11).

While there are no approved vaccines in the US, the attenuated ‘live vaccine strain’ (LVS) for tularemia has been used experimentally in the US since the 1950s (12). By further attenuating original tularemia vaccine stocks provided by the N. F. Gamaleya Federal Research Center for Epidemiology & Microbiology (13, 14), the US Army Medical Research Institute of Infectious Disease (USAMRIID) subsequently derived the LVS vaccine. The vaccine is derived from a milder subspecies of *F. tularensis* known as *Ft. holarctica* or type B, found in the Northern hemisphere and the only naturally occurring European subspecies. The vaccine has been used in nearly 4,000 volunteers since the late 1950s [See Table 1]. Both cellular and humoral immunity play a role in the vaccine’s effectiveness. It is administered by scarification, similar to the smallpox vaccine. The most reliable sign of successful immunization remains a ‘take’ reaction at the scarification site. This has been attributed to a delayed hypersensitivity T-cell response to the tularin protein (15). While the extent of clinical protective efficacy has been explored perhaps more than any other biodefense vaccine produced to date including human challenge trials, specific immune correlates of protection in humans remain elusive.

**Table 1:** Participant Demographics. Characteristics of participants screened for the two studies - FY 03-24 (2004-2008) and FY 07-15 (2009-2017).

| Study | FY 03-24 (N=484) |  | FY 07-15 (N=191) |  | Total |  |
| --- | --- | --- | --- | --- | --- | --- |
| Characteristic | N | % | N | % | N | % |
| **Sex** |  |  |  |  |  |  |
| Male | 310 | 64.0 | 112 | 58.6 | 422 | 62.5 |
| Female | 174 | 36.0 | 79 | 41.4 | 253 | 37.5 |
| **Race** |  |  |  |  |  |  |
| White | 421 | 87.0 | 162 | 84.8 | 583 | 86.4 |
| African American | 26 | 5.4 | 10 | 5.2 | 36 | 5.3 |
| American Native | 2 | 0.4 | 1 | 0.5 | 3 | 0.4 |
| Asian | 14 | 2.9 | 13 | 6.8 | 27 | 4.0 |
| Pacific Islander | 2 | 0.4 | 0 | 0 | 2 | 0.3 |
| Other | 19 | 3.9 | 5 | 2.6 | 24 | 3.6 |
| **Age** |  |  |  |  |  |  |
| < 20 | 2 | 0.4 | 0 | 0 | 2 | 0.3 |
| 20 - 29 | 128 | 26.4 | 53 | 27.7 | 181 | 26.8 |
| 30 - 39 | 152 | 31.4 | 77 | 40.3 | 229 | 33.9 |
| 40 - 49 | 124 | 25.6 | 46 | 24.1 | 170 | 25.2 |
| 50 - 59 | 66 | 13.6 | 14 | 7.3 | 80 | 11.9 |
| 60 - 69 | 12 | 2.5 | 1 | 0.5 | 13 | 1.9 |
| **Baseline Titer** |  |  |  |  |  |  |
| ≥ 1:20 | 49 | 10.1 | 54 | 28.3 | 103 | 15.3 |
| < 1:20 | 435 | 89.9 | 137 | 71.7 | 572 | 84.7 |

Although studied under IND for decades, LVS has never received marketing approval by a stringent regulatory authority. Pursuit of full licensure has been hampered by concerns regarding vaccine strain reversion to wild type (though this has not been documented in clinical studies), purported challenges for vaccine manufacture at commercial scale, and lack of a commercial partner. In the absence of an alternative candidate in clinical development, USAMRIID has continued to vaccinate laboratory workers at risk for occupational tularemia exposure under the Special Immunizations Program (SIP) (16). The SIP was maintained for several decades at USAMRIID and vaccinated workers from numerous bio-containment laboratories in the US working on tularemia and other select agents. The work has continued under IND in the hopes of adding to the clinical database, providing critical comparative data for emerging alternatives. The vaccine is also thought to impart volunteer laboratory workers a potential added layer of protection against *Ft* infection beyond standard biosafety laboratory practices. We report here data from two longitudinal protocols of LVS administered open label to at-risk personnel. The first study (FY03-24) began in 2004 and the second (FY07-15) concluded in 2017. At present no further studies of LVS are planned and these may represent the last clinical trial data of the LVS vaccine.

## Methods

### Study Design

Two consecutive longitudinal uncontrolled open-label, Phase 2 safety and immunogenicity studies of the LVS vaccine were conducted at USAMRIID under separate but similar protocols between 2004 and 2017 under IND 157 (US FDA). Individuals who entered containment laboratory suites where tularemia might be aerosolized, those with possible contact with contaminated equipment or materials, and in some cases those travelling to endemic countries were vaccinated. Placebo and/or other controls were not used as the volunteers were all at -risk of occupational laboratory-acquired tularemia infection.

### Ethics Statement

All human subjects research studies described were approved by the Institutional Review Board of the U.S. Army Medical Research and Development Command (formerly Medical Research and Materiel Command) prior to commencement. Protocol FY03-24 was conducted from 1 October 2004 to 6 October 2009, while protocol FY07-15 ran from 28 August 2009 to 4 December 2017. The studies were registered at ClinicalTrials.gov as NCT00584844 (trial FY03-24) and NCT00787826 (trial FY07-15). An additional protocol FY15-14 was registered as NCT03867162 but never enrolled subjects. Written informed consent was obtained from all subjects prior to enrollment.

### Vaccine

The vaccine used for both studies was *F. tularensis* Vaccine, Live, NDBR 101, Lot 4 manufactured by the National Drug Company in 1962 under Investigational New Drug Application 157. The vaccine consists of live, attenuated *F. tularensis*, in a modified casein partial hydrolysate medium manufactured on 28 May 1962. Vaccine was stabilized with a solution of glucose cysteine hemin agar and sucrose gelatin agar stabilizer solution without preservative and stored between −10° and −30° C. Potency testing in guinea pigs under Good Laboratory Practices (GLP) was performed on an annual basis by Southern Research Institute and/or USAMRIID and reported to FDA. Vaccine use was continued annually based on demonstrated protection of guinea pigs against challenge with virulent *F. tularensis* under FDA Good Laboratory Practices.

Vaccine vial vacuum seal integrity was confirmed using a high frequency generator (the SPARK test). Vials with an intact seal were reconstituted in 2.0 mL sterile water for injection, USP at concentration of approximately 1.0 × 10^9^ organisms/mL. Vaccine could be stored once reconstituted for up to 8 hours at 2-8° C. Roughly 0.06 mL of vaccine was administered to the volar forearm with 15 superficial punctures in protocol FY07-15, compared to 0.0025mL in protocol FY03-24. Excess vaccine was gently dabbed with sterile gauze and allowed to air dry for 30 minutes.

### Study Subjects

Volunteers at risk for infection with tularemia from various BSL-3 laboratories in the US were enrolled under USAMRIID IND protocols FY07-15 and FY03-24. Volunteers had to be at least 18-65 years old with known risk for *F. tularensis* exposure and willing to participate in the protocol for the duration of the study, approximately 6 months. They had to be medically cleared to participate by an investigator based on history, physical exam and laboratory testing. They could not have had a documented tularemia infection, been vaccinated against tularemia in the preceding 10 years for enrollment in FY07-15. For study FY03-24, volunteers could not have ever received a tularemia vaccination. Subjects could not have received antibiotics within the prior 7 days or vaccination with another vaccine within 4 weeks, be immunodeficient, have confirmed HIV or use immunosuppressing medications. Volunteers were also excluded if pregnant or lactating, allergic to vaccine components, or had clinically significant abnormal laboratory results within 60 days prior to vaccination.

### Experimental Methods

Following informed consent, volunteers were screened for inclusion in the study by medical history, physical examination, chest X-ray, electrocardiogram (EKG), CBC, chemistry, urinalysis, liver function tests, hepatitis and HIV antibodies. A baseline microagglutination titer was obtained and volunteers were excluded if positive. Females had a urine or serum pregnancy test. Those eligible were administered vaccine on study day 0, with follow-up for take reaction on days 1, 2 and between day 5 and 9. Volunteers returned between days 12-16 and 28-35 for adverse event assessment, with microagglutination titers (MA) obtained between days 28-35. A close-out interview, and final scarification site assessment were completed at 6 months for protocol 07-15 and at one year for 03-24. Second doses of vaccine were not routinely administered unless a subject did not have an initial take reaction. Subjects with insufficient initial responses to vaccine at 28-35 days had MA titers recorded. In protocol 07-15, subjects returned for follow-up examination on days 1 and 2, between days 5-9, 12-16, and 28-35, and at 6 months (±14 days) after vaccination or revaccination for clinical evaluation of AEs and to document responses to vaccine. Additionally, subjects were in some cases asked to return on days 56-84 for laboratory tests.

### Immunogenicity and Serology

Vaccination effectiveness was assessed based on formation of an erythematous papule, vesicle, and/or eschar with or without underlying induration, known as a ‘take reaction’ between 5 and 9 days after vaccination. Microagglutination titers (MA) were performed by the USAMRIID Research Serology lab using previously described methods (17, 18). Briefly, patient serum collected at the indicated time points below was incubated with a standardized 1:5 dilution *Ft* antigen for 24+/-4 hours at serial dilutions from 1:10 to 1:20,480. The highest serum dilution observed with an even lattice of agglutination and only a small transparent button of stained cells at the bottom of the microtube well was considered the endpoint titer. Rabbit antiserum was used as a positive control, and pre-tested human serum as a negative control. An MA titer >1:20 was considered a positive response in protocol FY03-24 while a 4-fold rise in titer from baseline were considered a positive antibody response for protocol FY07-15. The change in protocol FY07-15 was made at least partly in response to a participant who had a high background titer of 1:40. The participant was not vaccinated and subsequently developed tularemia due to a laboratory exposure(19). MA titer was repeated at day 56-84 if the day 28-35 MA titer increased less than 4-fold. If there was no positive take reaction following the initial vaccination, a second dose was recommended and offered at least 28 days after the previous vaccination. If there was no positive take reaction 28-35 days after the second dose, or MA titer increased less than 4-fold, a third dose was considered.

### Safety

Participants were monitored continuously for adverse events throughout the study. Adverse events were described using contemporary versions of the National Cancer Institute Common Terminology Criteria for Adverse Events (CTCAE). Adverse events and subject safety concerns were reviewed by the Investigator as well as the Sponsor’s clinical research and medical monitors. Subjects were monitored for 30 minutes following vaccination. Subjects were seen in clinic for follow-up on Days 1, 2, 7, 14 and 28. Study staff contacted subjects at week 3 by phone or email to ensure attendance at the week 4-5 safety and immunogenicity assessment.

### Statistical Analysis

At the conclusion of the study, data were transformed to SAS® data format using DBMS/COPY™ or system-specific data transfer programs. Descriptive tables of demographics, compliance, frequency and rate of each reaction, responder and non-responder rates, study deviations, concomitant medications, any local injection measurements or vitals taken, and out-of-reference range laboratory values were compiled.

Endpoint measurements were evaluated for all intent-to-treat subjects regardless of compliance with titer schedule. However, titers obtained on samples collected outside the protocol prescribed window were excluded from further analysis. Data from the final statistical analysis report from each study was reviewed. Key immunogenicity outcome measures included ‘take reaction’ and MA titers. MA titer criteria differed by study with a >1:20 considered positive in the FY03-24 study compared to a 4-fold rise for the FY07-15 study. Safety endpoints included vaccine relatedness of adverse events as well as frequency and severity.

For the purposes of this analysis comparisons of titer response and take reaction by categorical variables (i.e. sex) were conducted using t-test and Pearson’s chi-squared analysis respectively. Comparisons of titer response and take reaction by continuous variables (i.e. age) were conducted using simple linear regression and simple logistic regression analysis respectively. Statistical significance was defined at the two-tailed 95% confidence level. All analysis was performed using GraphPad Prism version 8.0.0 for Windows (GraphPad Software, San Diego, California USA, www.graphpad.com).

## Results

There were 484 subjects screened for study FY03-24 which ran between October 2004 and September 2008. Of the 462 vaccinated, 405 completed the final study visit at 6 months. To enroll in the study, subjects had to have a titer <1:20, no previous vaccination, and no documented tularemia infection. A small number (21) were revaccinated with the same initial dose of vaccine, based on inadequate response to the initial vaccination (titer <1:20) and two required a second revaccination (3^rd^ dose) (**Figure 1a**). There were 191 subjects screened and 177 vaccinated for study FY07-15 which ran between August 2009 and May 2017. In contrast to FY03-24, subjects could not have had a prior tularemia vaccination within the past 10 years. In FY07-15 subjects with inadequate titers were not revaccinated. Adequate titers were also defined differently as greater than 4-fold rise over baseline. There were 170 completing FY07-15 with 7 withdrawals before the final study visit at 6 months.

**Figure 1a:**
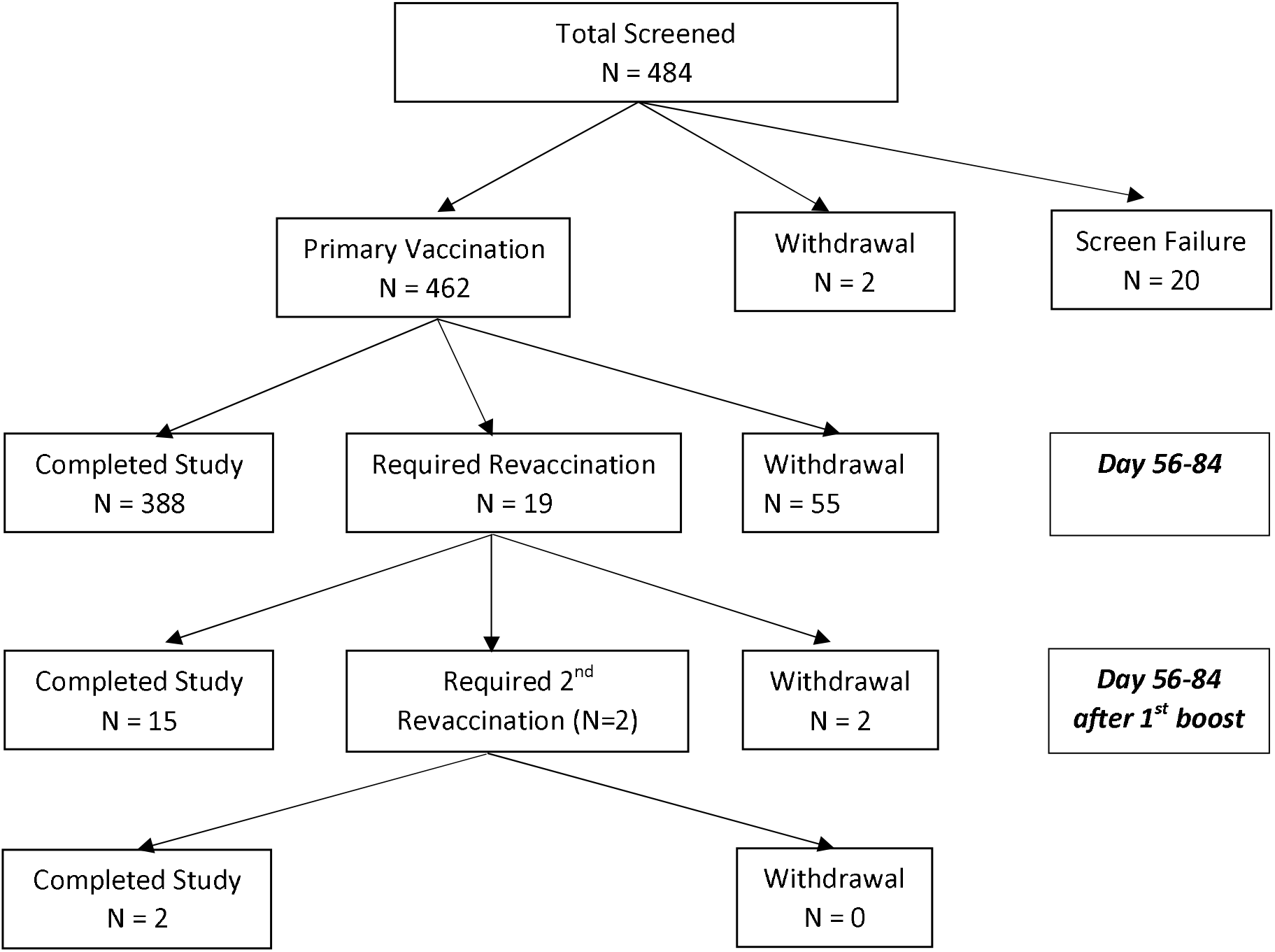
CONSORT diagram. Disposition of subjects enrolled to LVS tularemia vaccine study FY03-24 (enrollment from 10/01/2004 to 09/15/2008). Reasons for screening failures included having previously received the vaccine or having a positive titer (n=10), previous exposure (1), medical reason (1), not able to travel to USAMRIID to receive the vaccine (1), or no longer at risk (7).

**Figure 1b:**
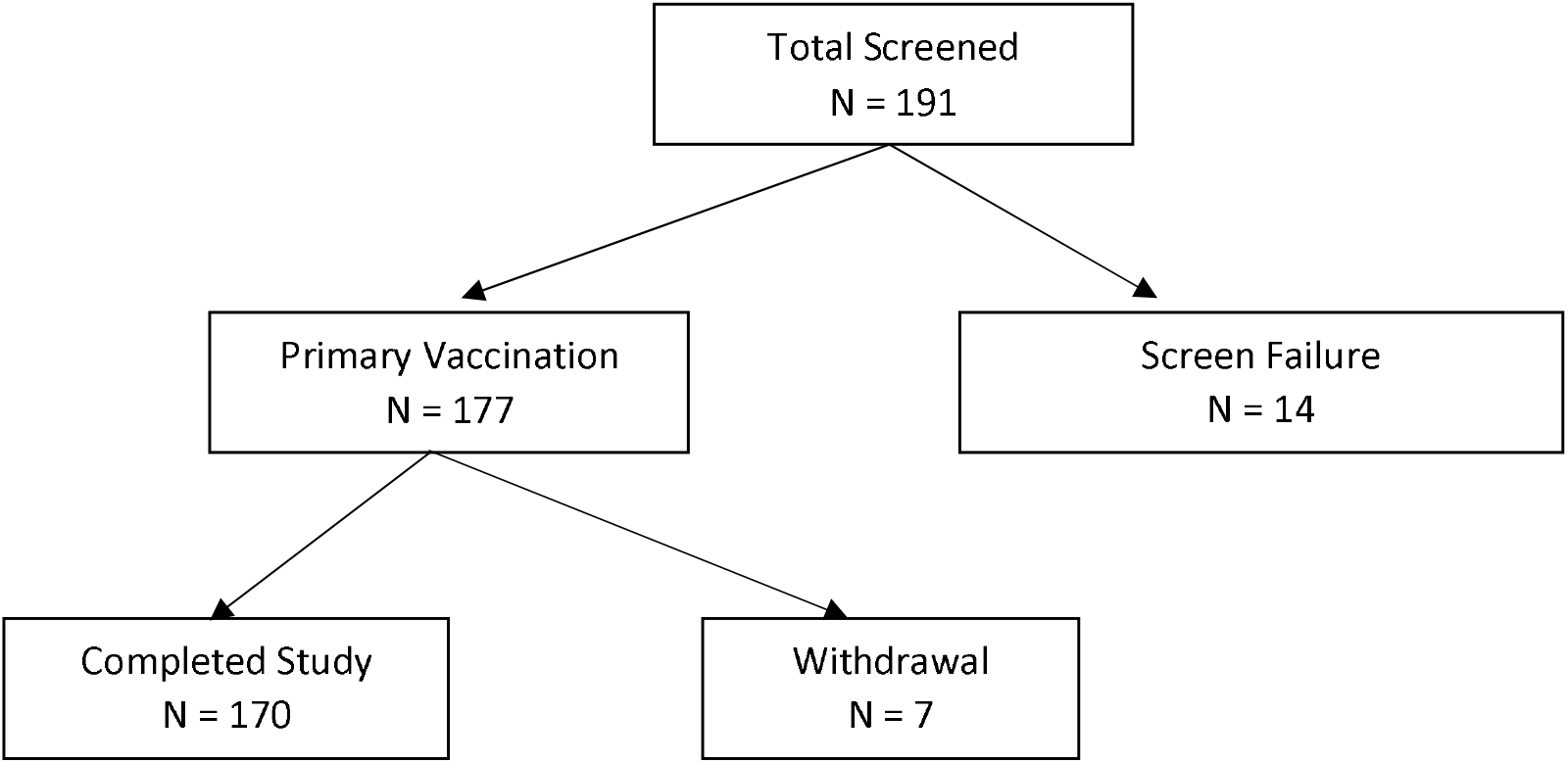
CONSORT diagram. Disposition of Subjects enrolled to LVS tularemia vaccine FY 07-15 (enrollment from 08/28/2009 to 05/05/2017). To enroll in the study, subjects could not have had a prior tularemia vaccination within the past 10 years. Reasons for screening failure included medical (5), relocation prior to vaccination (3), and no longer at occupational risk (6). Subjects were considered to be fully vaccinated if they had a 4-fold rise from baseline, and a positive ‘take reaction’.

Demographic characteristics of participants in both studies are shown in Table 1. More than 10% of subjects entering FY03-24, and 25% of those in FY07-15 had baseline microagglutination titers against tularemia >1:20. It should be noted that having a titer >1:20 was not exclusionary in FY07-15. More than 60% of subjects in both studies were 20-40 years old, >60% male, and >85% Caucasian.

### Immunogenicity

Overall, the Live *F. tularensis* Vaccine (NDBR-101, Lot 4) had comparable rates of immunogenicity to those seen in prior studies based on the two principle measures – microagglutination antibody titers and take reaction (**Table 3**). Immunogenicity response rates were uniformly high. In both studies, all protocol-compliant subjects had a measurable immune response (titer ≥ 1:20 and ‘take reaction’). A positive ‘take reaction’ was defined as development of one of the following: erythematous papule, vesicle, and/or eschar formation. All 177 subjects vaccinated (100%) in FY07-15 had a positive ‘take reaction’ within 9 days of vaccination, suggesting a robust qualitative cellular immunity response within 14 days. In FY03-24, 95.5% of subjects (n=441; SEM=0.010) had a positive take reaction at first vaccination. Subjects in FY03-24 without adequate initial responses were revaccinated. Positive take reactions were seen in 89.5% (n=17; SEM 0.007) of those receiving a first revaccination, and 100% receiving a 2nd revaccination. No statistically significant differences in ‘take reaction’ rates for any vaccination event were observed when analyzed by sex (via Pearson’s chi-square) or age (simple logistic regression).

In protocol FY03-24, of 454 protocol-compliant post-vaccination titers collected at day 28-35, 442 (98.6%) were ≥ 1:20. LVS was also highly immunogenic in the FY07-15 study where immunogenicity was defined as ≥ 4-fold rise in MA antibody titer following vaccination. This immunogenicity endpoint definition differed from prior studies, including FY03-24 where a 1:20 titer value was considered immunogenic. All but 9 subjects vaccinated in FY07-15 had a ≥ 4-fold rise in titer by day 28-35 and were not evaluated again. There were 9 subjects (5%) who had titers repeated at day 56-84, and of these 6 of 9 (67%) had a ≥ 4-fold rise in titer while 3 remained non-responders (33.3%). However, all 3 of the latter individuals had ‘take reactions’. One of the 3 non-responders developed an upper respiratory infection and was treated with antibiotics within the 1st week after vaccination, providing a likely cause for non-response. The study duration of individual subjects in the FY07-15 protocol was limited to 6 months.

Microagglutination titers over time are shown in **Figure 2A**. For study FY07-15 titers were obtained in 191 subjects at baseline (mean (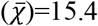), SEM=3.7), and 177 subjects at day 28-35 (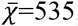, SEM=51.2). A small number (n=14) had follow-up titers at 6 months (µ=262, SEM=97.0). Titers for subjects from FY03-24 are shown at baseline (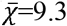, SEM=0.4; n=484), day 28-35 (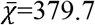=379.7, SEM=27.3; n=459) and 12 months (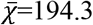, SEM=11.3; n = 407) after first vaccination. Responses were modest but nearly all >1:20 for the 19 subjects receiving a 1^st^ revaccination (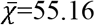, SEM=17.06), and 2 receiving a 2^nd^ revaccination (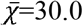, SEM=10). Geometric mean titers are shown in Figure 2B with baseline values for FY03-24 of 7.98 (95% confidence interval (95%CI) =7.69-8.28), and 9.56 (95%CI=8.67-10.54) for FY07-15. Peak geomean titers at vaccination day 28-35 were 181 for FY03-24 (95%CI=161–204) but significantly higher at 269 for FY07-15 (95%CI=223–323; p=0.0006 for t-test of log-transformed data). Month 12 geomean titers were 117 (95%CI=105–130) in FY03-24 while month 6 geomean titers where available for FY07-15 were 101.7 (95%CI=24.6–421).

**Figure 2.**
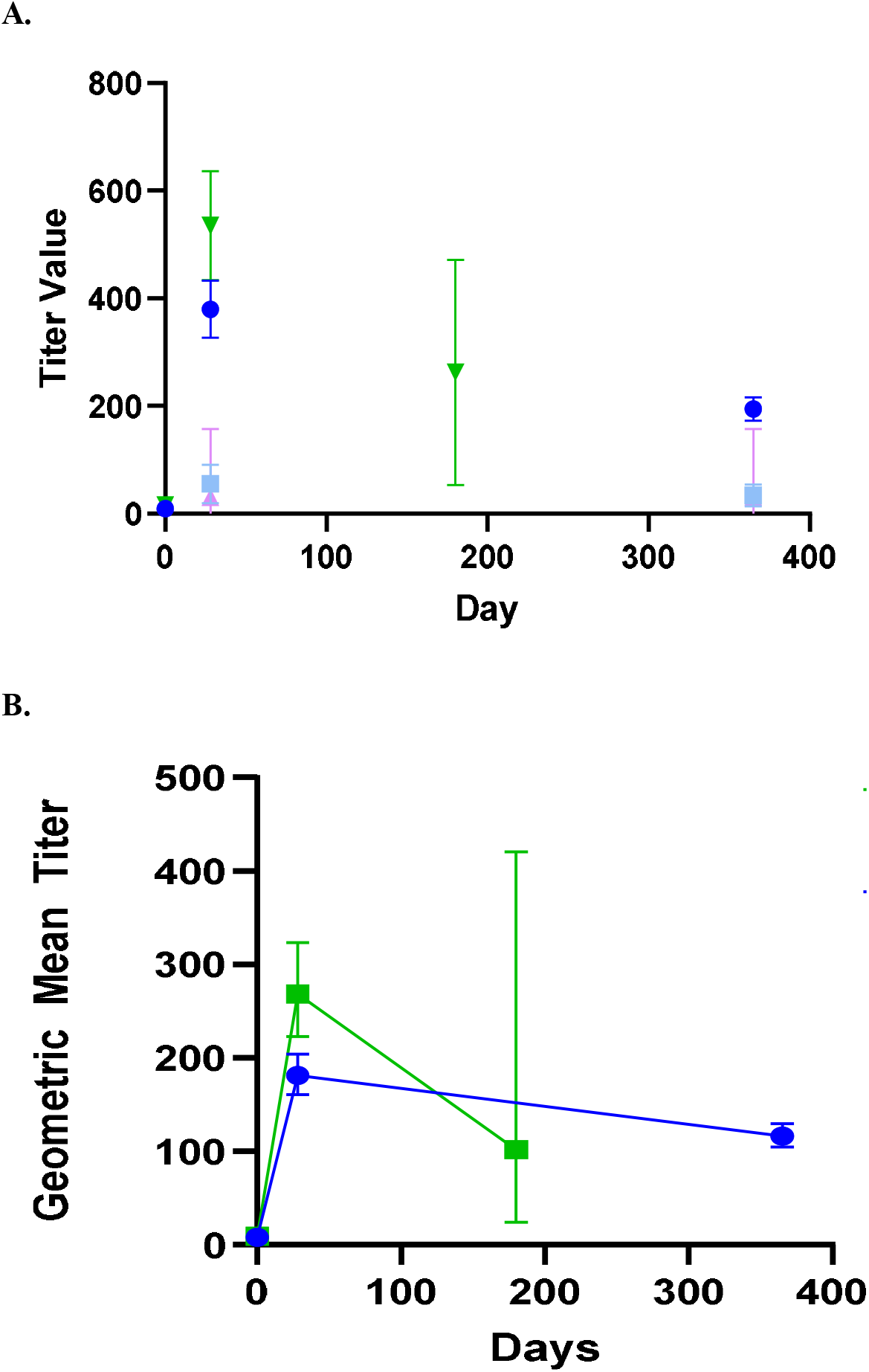
Microagglutination titers over time by study. **A**. Mean and standard error of the mean (SEM) bars for titer values are shown for the two studies. Titers were obtained at baseline, day 28-35, and 6 months for study FY07-15 (green triangles). All 177 subjects vaccinated (100%) in FY07-15 had a positive ‘take reaction’ within 14 days. Titers for subjects from FY03-24 are shown at baseline, and days 28-35 after first vaccination, and 12 months (blue circles). Mean and standard error bars are also shown for the 21 subjects receiving a 1^st^ revaccination (light blue squares), and 2 receiving a 2^nd^ revaccination (pink triangles). In FY03-24, 95.5% of subjects had a positive take reaction at first vaccination. 89.5% of those receiving a first revaccination, and 100% receiving 2^nd^ revaccination had a positive ‘take reaction’. **B**. Geometric mean titers with 95% confidence intervals for each time point are shown with green squares representing FY07-15 titers following initial vaccinations only, and blue circles representing FY03-24.

There were very few follow-up titers at the 6 month time point for FY 07-15 (n=14 of the original 191 vaccinated) as these were only performed if the day 28-35 response was inadequate. Substantially more had 12 month follow-up titers in FY 03-24 (n=407 of the 459 vaccinated). Where available, titer levels were largely sustained at month 12 for the small number revaccinated with mean values of 33.9 (SEM=9.7; n=18) for first revaccination and 30.0 (SEM=10) for the 2 undergoing a 2^nd^ revaccination. It is noteworthy that the same lot of vaccine was used for both studies, though titer responses were higher following initial vaccination in study FY07-15 (535 vs 380; p= 0.004).

There were no significant difference in mean titer values for either study by sex (**Figure 3A**). There was not a statistically significant difference between males and females for month 12 titer in FY03-24 (185.2 vs 209.8; p = 0.296). There was not a statistically significant difference by age for month 12 titer, though the observed modest decrease in titer with age did approach statistical significance (y=-2.081x+273.2; p = 0.0585). Simple linear regression of titer values by age revealed modest statistically significant declines with increasing age (**Figure 3B**) for study FY03-24 where there were more than twice as many subjects enrolled but not FY07-15.

**Figure 3.**
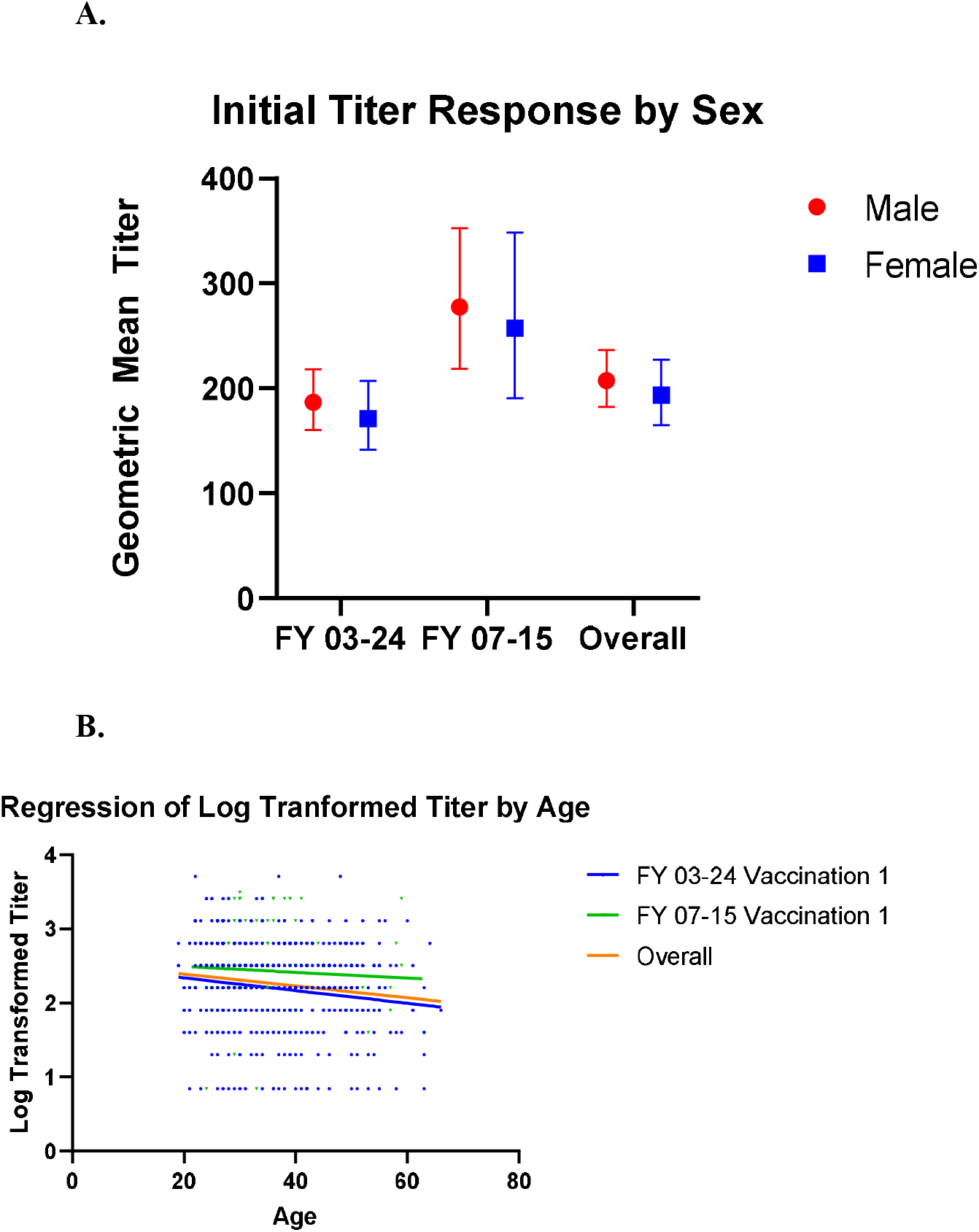
Microagglutination Titer Values by Age and Sex at day 28-35. A. Geometric mean microagglutination titer values are shown for male (red circle) and female (blue squares) subjects from Study FY07-15 and FY03-24 as well as for the two studies combined following first vaccination. There were no significant differences in either study by sex. Geometric mean titers (with 95% confidence intervals) in FY03-24 were 186.9 (160.2–218.1) for males and 171.2 (141.6 – 207.1) for females (unpaired t-test p = 0.488). Geometric mean FY07-15 titers were 277.6 (218.4–352.7) for males and 257.6 (190.5–348.5) for females (unpaired t-test p = 0.699). Overall geometric mean titers were 207.6 (182.2– 236.5) for males and 193.6 (164.7 – 227.5) for females (t-test p=0.5110). **B**. Simple linear regression of log-transformed titer values by age is shown for FY03-24 (blue lines/dots), FY07-15 (green lines/dots), and the two studies combined (orange lines). While there was a statistically significant decline by age in FY 03-24 (y=-0.00857x+2.510; p = 0.0016) and overall (Y = -0.00799 + 2.550; p = 0.0008), decline was not significant in FY07-15 alone (Y = -0.00392 + 2.571; p = 0.4088).

Adverse events are shown in **Table 2**. In study FY03-24, 401 of 462 vaccinated subjects (86.6%) had at least one adverse event during the course of the study. In FY07-15, 170 of 177 (96.1%) reported at least one AE. Local AEs were most common, reported in 84.1% in FY03-24, and 81.4% in FY07-15 with only 1 local event graded as severe in the former study. Scarification site lesions and pain were the most prominent overall, accounting for nearly 2/3 of local reactions. Systemic adverse events occurred in fewer than 17% of those vaccinated with leading causes including fatigue (4.8%), headache (3.9%), myalgia (2.9%) or fever/chills (1.5%). Pharyngitis, often unilateral and with a tonsillar exudate was seen in less than 1% of subjects. Most AE rates were consistent across the two studies, with the exception of lymphadenopathy which was noticeably higher in FY03-24 (18.2% vs 2.8%). Anatomic sites of observed lymphadenopathy in study FY03-24 included 65 axillary, 15 epitrochlear, 2 cervical, and 143 unspecified with 17 reported as epitrochlear in FY07-15 (and the remainder unspecified). Scarification site pain was more commonly reported in FY07-15 (43.5% vs 16.6%).

**Table 2:**
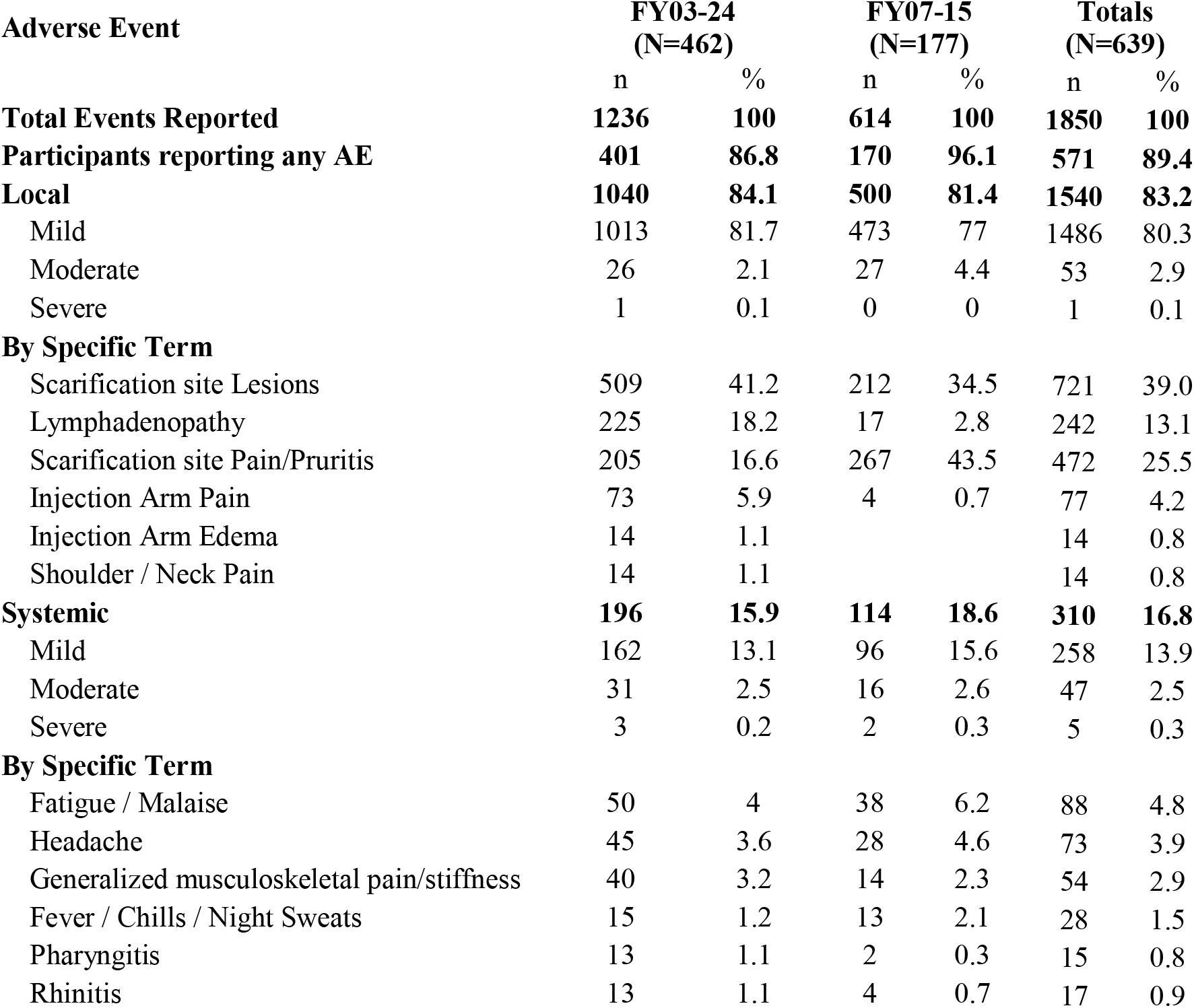
Adverse events reported following LVS vaccination. Adverse events reported at any time during the study period are shown. Only AEs determined by an investigator to be probably or definitely related to vaccination are included.

| Adverse Event | FY03-24<br>(N=462) |  | FY07-15<br>(N=177) |  | Totals<br>(N=639) |  |
| --- | --- | --- | --- | --- | --- | --- |
|  | n | % | n | % | n | % |
| **Total Events Reported** | **1236** | **100** | **614** | **100** | **1850** | **100** |
| **Participants reporting any AE** | **401** | **86.8** | **170** | **96.1** | **571** | **89.4** |
| **Local** | **1040** | **84.1** | **500** | **81.4** | **1540** | **83.2** |
| Mild | 1013 | 81.7 | 473 | 77 | 1486 | 80.3 |
| Moderate | 26 | 2.1 | 27 | 4.4 | 53 | 2.9 |
| Severe | 1 | 0.1 | 0 | 0 | 1 | 0.1 |
| **By Specific Term** |  |  |  |  |  |  |
| Scarification site Lesions | 509 | 41.2 | 212 | 34.5 | 721 | 39.0 |
| Lymphadenopathy | 225 | 18.2 | 17 | 2.8 | 242 | 13.1 |
| Scarification site Pain/Pruritis | 205 | 16.6 | 267 | 43.5 | 472 | 25.5 |
| Injection Arm Pain | 73 | 5.9 | 4 | 0.7 | 77 | 4.2 |
| Injection Arm Edema | 14 | 1.1 |  |  | 14 | 0.8 |
| Shoulder / Neck Pain | 14 | 1.1 |  |  | 14 | 0.8 |
| **Systemic** | **196** | **15.9** | **114** | **18.6** | **310** | **16.8** |
| Mild | 162 | 13.1 | 96 | 15.6 | 258 | 13.9 |
| Moderate | 31 | 2.5 | 16 | 2.6 | 47 | 2.5 |
| Severe | 3 | 0.2 | 2 | 0.3 | 5 | 0.3 |
| **By Specific Term** |  |  |  |  |  |  |
| Fatigue / Malaise | 50 | 4 | 38 | 6.2 | 88 | 4.8 |
| Headache | 45 | 3.6 | 28 | 4.6 | 73 | 3.9 |
| Generalized musculoskeletal pain/stiffness | 40 | 3.2 | 14 | 2.3 | 54 | 2.9 |
| Fever / Chills / Night Sweats | 15 | 1.2 | 13 | 2.1 | 28 | 1.5 |
| Pharyngitis | 13 | 1.1 | 2 | 0.3 | 15 | 0.8 |
| Rhinitis | 13 | 1.1 | 4 | 0.7 | 17 | 0.9 |

All adverse events, including serious adverse events, had onset dates up to 37 days post-vaccination. Duration of adverse events, including serious adverse events, ranged from 1 to 367 days. Two serious adverse events (SAEs) were reported. Both required hospitalization. One - a case of acute appendicitis 23 days after vaccination - resolved within 2 days. The second was a coronary artery disease event occurring 24 days after vaccine administration, which resolved in 21 days. Neither was determined to be related to the vaccine, nor deemed to be life threatening. **Figure 4** shows representative take reactions in 3 individuals from the FY03-24 study to illustrate typical appearance on the volar forearm.

**Figure 4.**
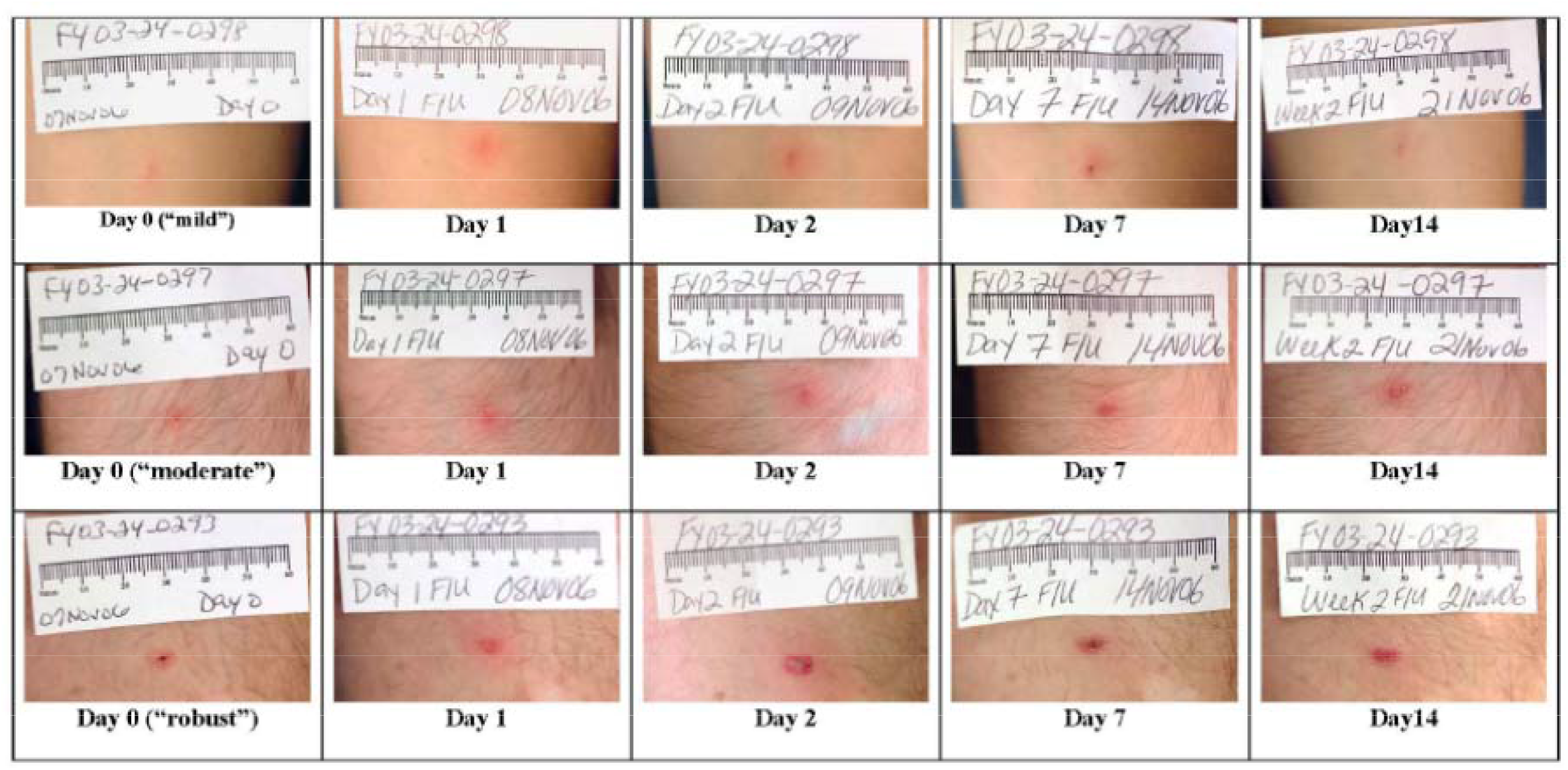
LVS live, attenuated tularemia vaccine “take reactions” in three representative individuals. A single dose of 0.06 ml of LVS vaccine at a concentration of 1.0 × 10^9^ organisms/mL was inoculated into the volar forearm of each volunteer using the scarification method. Although the three individuals depicted were immunized using the same technique, these pictures show a range of possible reactions. The three individuals had day 28-35 MA titers of 160, 80, and 640 respectively. As shown in the photographs, in all three cases there is appreciable healing of the site by day 14 following vaccination. By day 28 only a small hyper-pigmented macule remained at each vaccination site (not shown).

## Discussion

The results seen from the most recent LVS protocols at USAMRIID parallel those from a large body of clinical trials conducted from the mid-1950s to the present in late 2017. The data presented here from 2004 - 2017 may represent the final LVS doses provided to volunteers under an IND clinical study. The clinical experience with the *F. tularensis* Vaccine, Live, NDBR 101 and related LVS vaccine lots is summarized in **Table 4** below. Lot 4, NDBR 101 manufactured in 1962 has been in at least 1,998 volunteers under IND 157. Combined with literature reports using other vaccine lots, LVS has been administered to roughly 4,279 volunteers. Overall, the vaccine has been shown to be safe. Common side effects include local lymphadenitis and scarification site pain in up to 80% within a few days of inoculation. Short-lived mild to moderate systemic symptoms (fever, malaise, myalgia) are also common in 15-20% within a few days to one week of inoculation. There have been only a handful of serious adverse events reported over this long period of use, none of which have so far been attributed to the vaccine. Particularly remarkable is that vaccine lots used in the present were up to 55 years old, retaining remarkable potency under controlled storage conditions. Immunogenicity as measured by the ‘take reaction’ has been consistently high with 95-100% of volunteers reacting after the first vaccination. Nearly all respond after a booster, even many years after lot manufacture. Likewise, immune reactions as measured by micro-agglutination titers greater than 1:20 are typically seen in 90% of recipients by 4 weeks and approaching 100% by 8 weeks. These simple, well-established findings should serve as benchmarks for future tularemia vaccine candidates.

**Table 3:** Summary of LVS vaccine studies conducted at USAMRIID and partnered facilities. Between the mid-1950s and 2017, the LVS tularemia vaccine was tested in numerous clinical trials to include 1) Healthy volunteer studies for safety and immunogenicity 2) Human challenge studies with tuleramia via the cuteneous and aerosol routes with and 3) without vaccination, in the latter case to better understand the natural history of human challenge with the Schu-S4 tularemia strain 4) studies in otherwise healthy laboratory workers potentially exposed to tularemia. Data is included from both the published literature and that collected in studies conducted under IND #157 for *Tularemia Vaccine, Live, Attenuated*.

| Years | Study Title | N | Immunogenicity | Safety |
| --- | --- | --- | --- | --- |
| **1) Safety and Immunogenicity Studies; N = 181 from IND; N = 2009 from literature** |  |  |  |  |
| 1991-1993 | Evaluation of the Safety and Immunogenicity of a Live Bacterial Tularemia Vaccine in Normal Adult Subjects (Protocol 91-9, Johns Hopkins University) | 94 | # (%) MA titer $\geq 1:20$ / Take reaction<br>Lot 2R-85: 43 (91.5) / 47 (100)<br>Lot 3-85: 46 (97.9) / 44 (93.6) | SAEs: none<br>AEs: 273 total AE reported (Lot 2R-85 – 144; Lot 3-85 - 129) |
| 1987-1991 | Evaluation of New Lots of a Tularemia Vaccine, Protocol A: Initial Assessment of the Safety of *F. tularensis* Vaccine, Live, TSI-GSD-213, Lot 1R-85 (Protocol 87-4, Log A-4706)<sup>a</sup> | 69<sup>a</sup> | % take reaction: not reported<br># (%) MA titer $>1:20$:<br>Vaccine 33 (97)<br>Placebo 3 (9) | SAEs: 1 (Hodgkin's lymphoma, preexisting); 7 graded severe (4 vaccine / 3 placebo)<br># (%) AEs: 34 (100) treated<br>32 (91.4) untreated |
| 1975 | Evaluation in Volunteers of the Active-Rosette-Forming Lymphocyte Test as an Assay for Previous Immunization to Tularemia (Protocol 75-2) | 11 | Test of Foshay skin test – 6/6 with prior vaccination positive; 5/5 vaccine naïve negative | SAEs: none<br>% AEs: none |
| 1965 | Metzger et al, 1965 (20); Bartelloni-1969 (3)<sup>b</sup> | 185 | # (%) take reaction: 181 (98)<br>Partial protection from aerosol challenge; greater at lower dose | SAEs: none<br>AEs: 30% lymphadenitis; systemic AEs not reported |
| 1964 | Evaluation of Attenuated Tularemia Vaccine (LVS) from National Drug Company (Protocol 65-4) | 7 | % take reaction: 100<br>% MA titer $>1:20$: 100 | SAEs: none<br>% AEs: 5/7 lymphadenopathy |
| 1958 | Hornick et al, 1958 (22)<sup>c</sup> | 24 | % take reaction: 100 (24/24) | % AEs: not reported |
| 1958 | Hornick et al, 1958 (6)<sup>c</sup> | ~1800 | % take reaction: not reported<br>% MA titer $>1:20$: not reported | data in 308 subjects indicated local reaction; 25% axillary adenitis; fever $<10\%$; headache 6.5%; myalgia 5%; flu-like symptoms 14% |
| **Challenge Studies; N = 130 from IND; 282 from literature** |  |  |  |  |
| 1967 | Evaluation of Storage Stability of Tularemia Vaccine, Live, Attenuated NDBR 101, Lot 4 (Protocol 68-4/68- | 20 | % take reaction: 100<br># (%) MA titer $\geq 1:20$: 20 (100) | SAE: none post-vaccine<br>Systemic AE: 13 post-vaccine |
|  | 4A) [Inpatient 5yr storage stability human challenge study]<sup>e</sup> |  |  |  |
| 1966 | Aerosolized LVS vaccine followed by challenge; Hornick and Eigelsbach, 1966 (22)<sup>d</sup> | 253 | # (%) MA titer ≥ 1:20: 97% by 3 weeks; cutaneous inoculation same rate but slower | AEs: At low dose ($10^4$ org) 30% had mild systemic AEs; High dose ($10^8$) - 90% systemic AEs |
| 1964 - 1965 | Evaluation of Attenuated Tularemia Vaccine (LVS) from National Drug Company (Protocol 65-8)<br>Evaluation of Storage Stability of Living Vaccine Strain (LVS) of *Pasteurella tularensis* (Protocol 65-13) and Evaluation of Metabolic Changes in Immunized Subjects Exposed to Infectious Doses of *P. tularensis* (65-13A) | 20 | % take reaction: 100<br>% MA titer > 1:20: 100% by week 4 for both lots<br>Subject requiring antibiotics low-dose (2.5k cfu) - 1 of 4<br>high dose (25k cfu) - 4 of 4<br>unvaccinated controls - 6 of 6 | SAEs: none after vaccination (65-8) or challenge<br>% AEs post-vaccine: 70% lymphadenopathy, no other conditions mentioned<br>AEs more common in high-dose challenge group |
| 1964 | Evaluation of Attenuated Tularemia Vaccine (LVS) from National Drug [Biologic Research] Company (Protocol 65-7) | 12 | % take reaction: 100<br>% MA titer > 1:20: 83% (10/12) by week 4 | SAEs: none<br>% AEs: 8% fever; 75% lymphadenopathy |
| 1962 | NDBR 101 (from Lots 1, 2, 3, 4, or 6) using the scarification technique<sup>f</sup> | 78 | # (%) take reaction: 77 (99)<br>% MA titer > 1:20: 68/71 (96) | % AEs: 36% (28/78) lymphadenitis; no systemic AEs |
| 1961 | Saslaw et al-1961 (56)<sup>g</sup><br>Saslaw and Carhart-1961 (57)<sup>g</sup> | 29 | MA titers peaked at 4-8 weeks | % AEs: 50% lymphadenopathy |
| **3) Challenge Only Studies; N = 10** |  |  |  |  |
| 1966 | Respiratory *Pasteurella (Francisella) tularensis* in Man (Protocol 67-1) | 10 | % take reaction: N/A<br>% MA titer > 1:20: 100% by week 4 | SAEs: none recorded<br>% AEs: 100% fever, HA, malaise; 9 AEs graded severe |
| **4) Clinical Studies (Single Arm Phase 2 in Laboratory Workers); N= 1,667** |  |  |  |  |
| 2009-2017 | A Longitudinal Phase 2 Study for the Continued Evaluation of the Safety and Immunogenicity of a Live *Francisella tularensis* Vaccine, NDBR 101, Lot 4 in Healthy Adults At-Risk for Exposure to *Francisella tularensis* (Protocol FY07-15) | 177 | # (%) take reaction: 177 (100)<br># (%) MA > 1:20: 165 (93.2) by d28; 171 (97.1) by d84 | # (%) SAE<sup>h</sup>: 2<br># (%) local AE: 173 (97.7)<br># (%) systemic AE:<br>Transaminase elevations: |
| 2004-2009 | A Longitudinal Phase 2 Study for the Continued Evaluation of the Safety and Immunogenicity of a Live *Francisella tularensis* Vaccine, NDBR 101, Lot 4 (Protocol FY03-24)<sup>i</sup> | 484 | # (%) take reaction: 441/462 (95.5) after primary dose<br># (%) MA titer > 1:20:<br>Primary - 442/454 (97.4)<br>Boost - 14/19 (73.7) | SAEs: none<br># (%) related AE: 399 (86.4)<br># (%) systemic AE: 264 (57.1) |
|  | No protocols conducted between 1999-2004 |  |  |  |
| 1987-1999 | Evaluation of New Lots of Tularemia Vaccine, Protocol B: Comparative Assessment of *F tularensis* Vaccine, Live, TSI-GSD-213 Lot 1R, and *F tularensis* Vaccine, Live, NDBR 101 Lot 11 (Protocol 87-9, conducted at USAMRIID and 18 extramural sites)<sup>j</sup> | 284 | Take reaction: not reported<br>#(%) MA >1:20: 152/224 (68.3) at 1 month; 208/224 (92.9) by 12 months | SAEs: none, 2 graded severe<br>AE data not recorded for 128 of 297 doses administered |
| 1965-1988 | Evaluation of Stability and Protective Efficacy of Tularemia Vaccine, Live, Attenuated, among At-Risk Personnel (Protocol AB-104)<sup>k</sup> | 722 | % take reaction: 100<br># (%) MA titer ≥1:20: 298 of 312 negative at baseline (99%) seroconverted | SAEs: none<br>% AEs: 87% had at least 1 AE; 90 AEs out of 722 subjects were considered related to vaccine |

**Table 4:** Protective effects of live attenuated tularemia vaccine (LVS) or prior tularemia exposure against subsequent tularemia challenge. Summary of historical literature where subjects underwent tularemia challenge after prior exposure or being randomized to vaccination with LVS or unvaccinated control groups. In A. subjects were vaccinated with LVS and then challenged by aerosol, while in B. they had been challenged previously without intervention, thus developing immunity to the disease. Subjects in C. were vaccinated with LVS, then challenged intradermally. In each experiment, subjects underwent subsequent challenge with the virulent SCHU-S4 strain at the doses and route indicated.

| A. LVS with aerosol challenge |  | # Ill vs. # Challenged |  | # Ill Requiring Treatment |  | Reference |
| --- | --- | --- | --- | --- | --- | --- |
| Challenge Dose (No. of Organisms) | Vaccinated | Control | Vaccinated | Control |  |  |
| 10 - 50 | 3/18 | 8/10 | -- | -- |  | Van Metre-1959 |
| 250 | 1/6 | 2/2 | 0 | 2 |  | Falbich-1946 |
| 2,500 | 2/5 | 2/2 | 0 | 2 |  | Falbich-1946 |
| 25,000 | 28/31 | 12/14 | 18 | 12 |  | Falbich-1946 |
| Total | 34/60 | 24/28 | 18 | 16 |  | Paviovskiy-1953 |

| B. Prior challenge with aerosol challenge |  | # Ill vs. # Challenged |  | # Ill Requiring Treatment |  |
| --- | --- | --- | --- | --- | --- |
| Challenge Dose (No. of Organisms) | Infected | Control | Infected | Control |  |
| 2,500 | 3/8 | 2/2 | 2/3 | 2/2 | McCrumb-1962 |
| 25,000 | 6/10 | 2/2 | 4/6 | 2/2 |  |
| Total | 9/18 | 4/4 | 6/9 | 4/4 |  |

| C. LVS with intra-dermal challenge |  | # Ill vs. # Challenged |  | # Ill Requiring Treatment |  |
| --- | --- | --- | --- | --- | --- |
| Challenge Dose (No. of Organisms) | Vaccinated | Control |  |  |  |
| 2,500 | 1/10 | 11/11 | Not reported |  | Sawyer -1962 |
| 8,200 | 1/8 | - |  |  |  |
| 100,000 | 1/3 | - |  |  |  |
| Total | 3/21 | 11/11 |  |  |  |

As a result of years of diligent effort, a tremendous amount is known about the clinical characteristics of live, attenuated self-replicating tularemia vaccines. While there has not been an opportunity to demonstrate vaccine efficacy against a deliberate biological attack, carefully conducted human challenge studies under Operation White Coat using the SCHU-S4 strain reveal important insights into vaccine performance parameters (10). Antibody levels > 1:20 by microagglutination assay (thought to be protective) were reached in the vast majority of volunteers following a single LVS dose (**Table 3**). **Table 4** summarizes published challenge study reports. LVS was protective in most cases against low-concentration SCHU-S4 challenge (2,500 organisms), though vaccine efficacy could be overcome at higher challenge concentrations (25,000 organisms) - 100-1,000-fold times greater than the minimum human infectious doses (3, 20, 21). Protection was afforded against both aerosol and intradermal challenge, though greater against the latter; **see Table 4**. While it is unlikely such studies could be approved today on ethical grounds, it should be noted that it was rare even for unvaccinated subjects challenged with low concentrations of tularemia to require antibiotic therapy. Many vaccinated subjects did not require antibiotic therapy at higher challenge concentrations, though unvaccinated subjects did so universally (22). In addition to direct evidence from challenge, indirect evidence comes from demonstrated reductions in occupational exposure infections among at-risk laboratory workers after introduction of LVS (23). The role of vaccination in an individual with an active infection or after virulent tularemia exposure is unclear, and should be evaluated in the future.

Microagglutination titers remained remarkably stable in the present study spanning 13 years of vaccination with the same product lot manufactured in 1962. The resiliency of present vaccine lot immunogenicity observed is consistent with a recent active comparator study. No differences were found in take reaction or seroconversion by microagglutination between the USAMRIID LVS vaccine and a newer vaccine lot manufactured for which NDBR 101, lot 4 served as seed material (24). Surprisingly, mean titers were modestly higher among subjects in the later study (FY07-15) despite the continued aging of lot 4. While there is no immediate explanation for this, titers were only modestly elevated (535) compared to averages in FY03-24 (mean=380). Despite this long term stability and the aforementioned protective efficacy, specific immune correlates of protection remain elusive. The role of humoral immunity in clinical protection from tularemia remains controversial and poorly understood. Conventional wisdom would indicate that an intracellular bacterium might require a cellular immune response (25), though an extracellular form of the organism has been identified (26). In humans, circulating antibodies to *F. tularensis* were initially measured with agglutination assays and enzyme-linked immunosorbent assays (ELISA) (27-29). In clinical research, a >1:20 microagglutination titer has been used to indicate an adequate vaccine response. LVS administered to subjects by scarification has reliably exceeded this benchmark over more than 6 decades (see **Table 3**). Bacterial agglutinins have been shown to increase and persist for at least 3 years (12). Limited evidence in mice indicates that antibody response is targeted to the lipopolysaccharide (*Ft* LPS) in both mice and humans. At least partial immunity is provided in passive antibody transfer studies from both murine and human donors with prior LVS vaccination (30, 31) as well as levofloxacin-treated mice who survived SCHU-S4 challenge (32). This appears to be mediated by the Fc-γ receptor (FcγR) (33). Currently available serologic markers of infection are not helpful for real-time diagnosis given a typical delayed rise in IgM, concurrent with IgG 2-3 weeks after symptom onset. Nor do they indicate successful treatment as IgG can remain elevated for several years (34). Recently, an outbred rabbit (Rb) model, coupled with a series of live attenuated S4-based vaccines (S4ΔaroD, S4ΔguaBA, and S4ΔclpB) and wild type Schu S4 aerosol challenge (∼2,000 CFU) were used to identify clinically measurable correlates of protection (COP). Analysis of individual rabbits (Rb) enabled retrospective identification of Rb COP that predicted S4 challenge outcome. In pilot studies, the group found several “Antigenic” COP in plasma of 80% of LVS-vaccinated humans (35).

It is thought that cellular immunity may play a dominant role in protection from tularemia given intracellular replication, predominantly in macrophages (36). The ‘take reaction’ following LVS vaccination is the most reliable indicator of cellular immunity, occurring in 90% or more of vaccinations (**Table 4**). However, take reaction assessment has been implicated in divergence between take and MA titer results, highlighting the importance of experienced readers. Immunogenicity in the present study differed from a recent study comparing USAMRIID LVS to a more recently manufactured batch by Dynport Vaccine Company (DVC-LVS) where there was greater discordance between MA and take reaction results (24). Following immunization or exposure to disease, a rarely used skin test (the Foshay test) has been shown to remain positive for up to 10 years (15, 37). Cellular immunity in humans is also measurable by a lymphocyte transformation test (38). While CD4 and CD8+ T-cells have been thought to be predominantly responsible, roles of dendritic cells, macrophages, neutrophils and natural killer cells have also been described (36). Cellular responses appear 2 weeks after vaccination as well as tularemia disease (38, 39), in comparison to serology which typically becomes positive in 2-4 weeks (40). However, evidence for the precise immune components involved in cell-mediated immunity remains incomplete. It appears likely both humoral and cellular immunity play a role. Passive transfer of spleen cells from vaccinated mice were protective against challenge with doses of LVS normally lethal to mice (but not humans) (41). Transfer of thoracic duct lymphocytes from immunized rats reduced the burden of infection following a low dose challenge with Ft LVS organisms in naïve rats, while rats given serum from immunized rats were not protected against Ft challenge (42). Rats vaccinated with aerosolized LVS survived subsequent lethal aerosol challenge with SCHU S4 without systemic infection. Tularemia replication in the lungs and characteristic pulmonary granulomatous lesions were reduced substantially though not eliminated (43), suggesting cellular protection. However, the translation of animal model findings to humans has remained elusive. At least some recent progress has been made at elucidating correlates in a rat co-culture model (44). In the model, T-cells from vaccinated rats are incubated *ex vivo* with tularemia-infected rat macrophages. It is thought that the same approach might be applied to human vaccine studies better elucidate cellular correlates of protection. A human model has been developed but has yet to be validated or submitted to a stringent regulatory authority for approval as a surrogate endpoint (45).

There may also be a role for mucosal immunity, given that both aerosolized LVS and aerosolized tularemia itself lead to development of immunity, the former more rapidly though of the same peak magnitude by 4 weeks as the intradermal route (22). When comparing cutaneous to aerosol vaccination in non-human primates, vaccine organisms were recoverable from the site of inoculation initially. Within 28 days they could be isolated from lymph nodes, liver and spleen but were cleared from the body by 90 days (12). IgA deficient mice were not protected against aerosol challenge following vaccination (46, 47). Immunity following aerosol challenge has been demonstrated in non-human primates (*Macaca mulatta*) with a clear dose-response enhancing survival in animals given higher LVS doses (10^7^ organisms) (22). In addition to dermal inoculation, LVS has proven to be protective as an aerosolized vaccine in humans, though side effects following higher doses required for protection may limit the practicality of this approach. In comparison, one study indicates that side effects at higher doses of inhaled SCHU S4 strain organisms (10^6^ – 10^8^) included fever and flu-like symptoms lasting 2-3 days (6). Syrjala *et al*. demonstrated that all of the 6 of 16 individuals protected against challenge with the virulent SCHU S4 *Ft*. strain by previous aerosol exposure to LVS had measurable circulating antibody while only one the 10 individuals not protected did (48). Of note, the individuals in this study had active tularemia infections, confirmed with serology, rather than vaccinated individuals. Whether LVS would be effective and marketable via trans-mucosal delivery route remains to be seen but may be worth investigating among vaccine candidates.

There is no identified replacement or competitor to LVS in clinical stage development at the time of writing. Key characteristics of an optimal tularemia vaccine could include lack of potential for reversion to wildtype tularemia in the case of live vaccines, a safety profile in humans equivalent or better than LVS, ease and low cost of manufacture, and efficacy against *Ft*-type A challenge in well-controlled large animal models. Comparable human immune correlates would need to be demonstrated in healthy volunteers, promising approaches demonstrated in murine (49) and human *ex vivo* co-culture model (45). A large slate of candidates have been produced over the years, and there is an excellent review on the subject published elsewhere (50). Approaches have focused on live, attenuated and inactivated tularemia vaccines employing various *Francisella* strains, particularly *F. holoarctica* and *F. novacida* (51). Inactivated vaccines may provide a safety advantage over live attenuated vaccines, but have yet to demonstrate comparable protection in animal models (46). Protein subunit vaccines would have a decided safety advantage, and also eliminate concerns for pathogenic strain reversion. However, this approach has yet to yield an effective candidate in animal models. DNA and vectored vaccine approaches using attenuated virus or bacteria for the purpose of introducing tularemia antigens have similarly resulted in suboptimal levels of animal protection (50, 52). Vaccines targeting the Fc-gamma receptor (FcγR) have been developed, given FcγR’s role in cellular phagocytosis and antigen presentation with mixed success (53).

In summary, the LVS vaccine was well tolerated and proved once again to deliver high levels of positive take reactions and microagglutination titers in at-risk laboratory workers over a 13 year period. Of particular interest is the resilience of the vaccine in long-term storage at USAMRIID. Despite manufacture 42-55 years prior to the studies, sustained high response levels continued to be observed. An incompletely understood mechanism of attenuation, hypothetical concerns for potential reversion to wildtype and instability in culture remain challenges for licensure and (54). A successful effort to remanufacture and test the vaccine in 2006 demonstrated that LVS could be manufactured using modern methods (24). LVS attenuation mechanisms have also been partially elucidated with mutations in a Type IV pilin gene (pilA) and an outer membrane protein (FTT0918) implicated in murine model virulence (55). Despite use for more than 70 years and a large safety database, there is currently no plan to pursue full licensure of LVS. As potentially the last laboratory worker was vaccinated under a clinical trial in 2017, the present work serves as a benchmark for safety, immunogenicity and long term potency against which to compare future alternative vaccine candidates. The long term potency of LVS would lend itself to a new vaccine protocol under IND for at risk personnel. These would reasonably include laboratory workers, military personnel and/or limited disaster preparedness stockpiles intended to deter deliberate biowarfare use. Given the US has no licensed measure for immunoprophylaxis against tularemia, a critical biodefense gap against a category A agent remains. Significant ongoing investment is needed.

## Data Availability

All data produced in the present work are contained in the manuscript.

## Acknowledgements

The authors would like to acknowledge the hard work and dedication of all of the members of the investigative and regulatory teams who supported tularemia protocols over the years at USAMRIID, particularly Mark Goldberg, Robert Rivard, Craig Koca, Jason Regules, Mark Paxton, Dixion Rwakasyaguri, Sandi Parriott, Lisa Borek, Paileen Mongeli, Latricia Dye, Cynthia Donovan, Denise Clizbe, Kimberly Pozzouli, Arthur Freedlander, Lawrence Korman, Vincent Fulton, Joel Bozue, and Beverly Foghtman. Most importantly we would like to acknowledge the volunteers who did the hard work to advance knowledge on countering tularemia and other biothreats by virtue of their professions and their selfless service.

